# Epidemic graph diagrams as analytics for epidemic control in the data-rich era

**DOI:** 10.1101/2022.10.10.22280897

**Authors:** Eugenio Valdano, Davide Colombi, Chiara Poletto, Vittoria Colizza

## Abstract

COVID-19 highlighted how modeling is an integral part of pandemic response. But it also exposed fundamental methodological challenges. As high-resolution data on disease progression, epidemic surveillance, and host behavior are now available, can models turn them into accurate epidemic estimates and reliable public health recommendations? Take the epidemic threshold, which estimates the potential for an infection to spread in a host population, quantifying epidemic risk throughout epidemic emergence, mitigation, and control. While models increasingly integrated realistic host contacts, no parallel development occurred with matching detail in disease progression and interventions. This narrowed the use of the epidemic threshold to oversimplified disease and control descriptions. Here, we introduce the epidemic graph diagrams (EGDs), novel representations to compute the epidemic threshold directly from arbitrarily complex data on contacts, disease and control efforts. We define a grammar of diagram operations to decompose, compare, simplify models, extracting new theoretical understanding and improving computational efficiency. We test EGDs on two public health challenges, influenza and sexually-transmitted infections, to (i) explain the emergence of resistant influenza variants in the 2007-2008 season, and (ii) demonstrate that neglecting non-infectious prodromic stages biases the predicted epidemic risk, compromising control. EGDs are however general, and increase the performance of mathematical modeling to respond to present and future public health challenges.

## Main text

Public health response to infectious disease epidemics faces two major challenges. First, to predict if an emerging pathogen will cause a large-scale epidemic. Second, to control epidemics and prevent recurring outbreaks of known pathogens. SARS-CoV-2 is evidence of both: a new pathogen which caught the world by surprise, wreaked havoc, and is now testing our control capabilities with recurrent waves [1]. Global crises like the COVID-19 pandemic may become more frequent, as climate change increases the threat of new viral species jumping to humans from animal reservoirs [2]. Among them, avian strains of influenza A (e.g. H5N1, H7N9) are closely monitored for their ability to develop into major pandemic strains [3]. With a universal vaccine still unavailable, preparing for the next influenza pandemic requires large-scale access to antiviral treatment [4] and optimal drugs administration [5, 6].

The fight against epidemics of known pathogens is equally hard, and exceptionally relevant in the context of sexually transmitted infections (STI). The progress towards elimination of HIV/AIDS is faltering [7]. Chlamydia, gonorrhea and syphilis cause more than 200 million new cases worldwide among adults each year [8], and widespread antimicrobial resistance is compromising our ability to fight them [9]. Yet elimination remains a priority, to reduce not only their burden, but also their effect on HIV acquisition and transmission [8]. Finally, monkeypox virus is currently spreading in non-endemic areas, overwhelmingly through sexual transmission and in communities already burdened by other STIs [10].

The two epidemic contexts – pandemics and STI – have little in common. But the goals of preventing large-scale emerging epidemics and eliminating endemic diseases find a common theoretical framework and application tool in the epidemic threshold [11, 12]. The epidemic threshold defines the critical value of disease transmissibility above which the infection can establish itself in a host population. Interventions that raise the epidemic threshold make the population more resilient to the pathogen. Opposite changes increase the epidemic risk.

The epidemic threshold has historically contributed to inform core public health activities, for its potential to provide context awareness and gauge the effort needed for prevention or eradication [13, 14, 15]. In the current COVID-19 pandemic, scientists, public health officials, authorities, and even the general public have continuously evaluated control policies by their ability to bring epidemic waves below the epidemic threshold, i.e., reproductive number *R*_*t*_ *<* 1 [16, 17, 18, 19].

Current theories, however, cannot handle the complexity of infectious disease dynam-ics in real populations, and resort to two oversimplified schemes. One consists in assuming that individuals mix at random, and stemmed from mathematical epidemiology [20]. This clashes, however, with the evidence of highly complex time-varying contact patterns measured or estimated in different contexts [15, 21, 22, 23]. To overcome this, the physics and network science community has pushed the development towards increasing realism in population structure [12, 24, 25, 26, 27, 28, 29]. This has happened, however, at the expense of disease description [30, 31, 32, 33], which has opened new problems: Simplifying assumptions on disease natural history bias results [34], and variations in intervention protocols cause radically different epidemic outcomes [6]. These simplifications limit our knowledge of disease dynamics, and cripple the ability of models to serve public health.

The new theoretical formalism presented here will eschew both. It will give estimates of the epidemic threshold that are both robust, to provide rapid and generalizable understanding of complex dynamical processes driving disease spread, and accurate, to turn high-precision data [35, 36, 37, 38] into targeted and viable recommendations. Applied to syphilis transmission, it shows that the resolution of sexual contact data requires matching resolution in modeling disease progression. Namely, its prodromic stage affects the epidemic threshold even if non-infectious. Applied to influenza, our framework predicts whether antiviral-resistant variants will become dominant, in agreement with the observed emergence and fixation of the oseltamivir-resistant A(H1N1) variant in 2007.

## Epidemic Graph Diagrams

We introduce here Epidemic Graph Diagrams (EGDs) to represent the coupled dynamics of arbitrarily complex disease spread, and arbitrarily complex contact patterns among hosts. Following a renowned tradition in physics [39, 40], our diagrams are more than a representation of the diffusion equations on the network substrate: they completely replace them. We define rules to build the diagrams, and operations to manipulate them, by exploiting their topological properties. Then, we show that diagrams lead to a general, analytical derivation of the epidemic threshold. Also, diagram operations simplify its numerical computation, making EGDs a practicable analytics for public health.

We consider the classic compartmental description of the natural history of an infectious disease, through a progression of successive disease states regulated by transition rates. We consider population structure through an explicit time-varying contact network with adjacency matrix *A*(*t*), where nodes represent hosts and the entry *A*^*ij*^(*t*) encodes transmission-relevant connectivity between host *i* and host *j* at time *t* [12, 15, 21, 22, 41].

Contacts can thus change in time.

The Markov chain formulation in the quenched mean-field approximation [24, 26, 28] of a generic compartmental model can be written as:

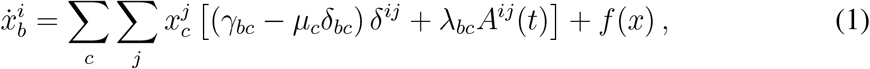

where 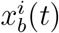 is the probability that node *i* is in compartment *b* at time *t*, and *b* runs over all compartments, except the susceptible S. *µ*_*c*_, *γ*_*bc*_, *λ*_*bc*_ are the rates of the transitions between compartments depicted in Fig. 1a: *µ* correspond to spontaneous transitions to S (e.g. recovery with no immunity); *γ* correspond to spontaneous transitions between any two compartments except S (e.g. hosts incubating the disease before becoming infectious); *λ* correspond to transmission events causing a susceptible node to enter another compartment (e.g. a susceptible becoming infectious). Transmissions are generated by infectious hosts of any kind (e.g. asymptomatic or symptomatic infectious individuals) when they are in contact with susceptible nodes. The component *f*(*x*) in Eq. (1) contains all terms that are nonlinear in *x* (quadratic or higher). It may be complex, but its derivatives vanish in the disease-free state 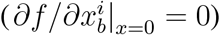, therefore *f* does not contribute to the epidemic threshold and can be dropped. Eq. (1) can then be conveniently rewritten in operator form:

**Figure 1.**
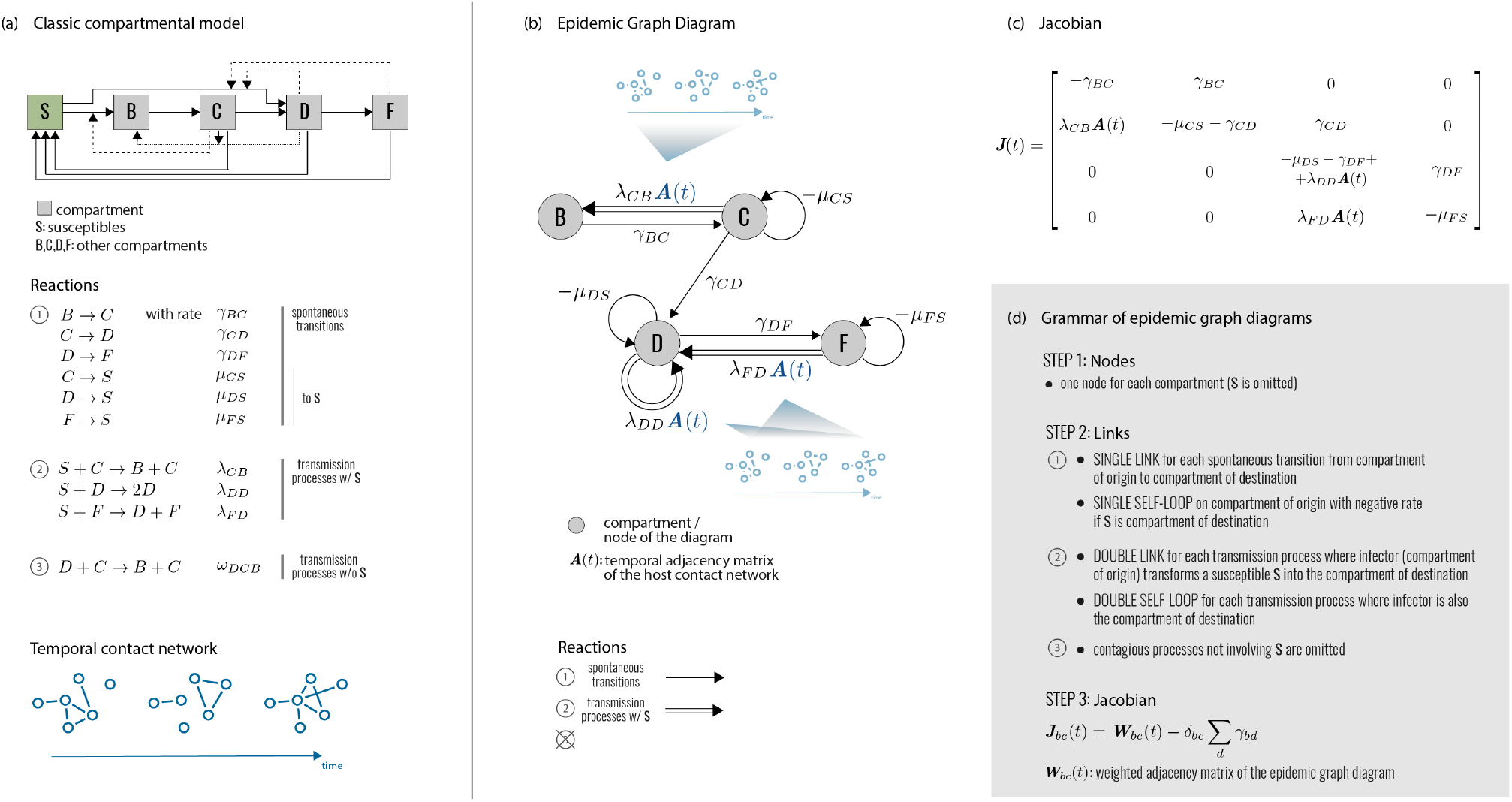
Epidemic Graph Diagrams. **(a)** Network epidemiology ingredients: compartmental model (top), and time-evolving network (bottom). The generic compartmental model appears in its standard representation: squares represent the different compartments, joined by transitions of three types. The susceptible compartment S (healthy individuals who can contract the disease from the infectious) is made explicit. Type 1 transitions are shown as continuous lines and correspond to spontaneous processes; type 2 are shown as dashed lines, for transmission events involving susceptibles; type 3 are shown as dotted lines, for transmission events not involving susceptibles. **(b)** Epidemic Graph Diagram corresponding to the compartmental model of panel a. Spontaneous transitions (type 1) become single links. Transmission events (type 2) become double links. Links are weighted by the corresponding transition rates (see panel a) multiplied by an operator on ℝ^*N*^. This operator is the identity matrix on single links (omitted), and the adjacency matrix on double links. All other transitions, e.g. transmissions infecting other compartments than S (type 3) can be neglected and do not appear in the EGD (see Methods). **(c)** Jacobian corresponding to the example in panels a, b. **(d)** Rules of the EGD grammar. The EGD is built directly from the compartmental model (panel a) following steps 1 and 2. The Jacobian is then the weighted adjacency matrix of the EGD minus diagonal terms that enforce the probability conservation (step 3), encoded in the diagonal entries *γ*_*bb*_. These are not free parameters but are fixed by probability conservation, since transitions among compartments not involving S do not change the sum 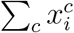 (Eq. 1). From this, we derive 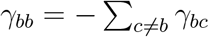.

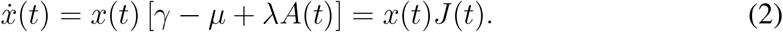

In this formalism, *x* is a vector on 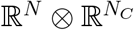, where *N* is the number of hosts, and *N*_*C*_ is the number of compartments (excluding S). *µ, γ, λ* are operators on 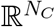, and *A* is an operator on ℝ^*N*^. The term *J*(*t*) = *γ* − *µ* + *λA*(*t*) is the Jacobian of the system, itself an operator on 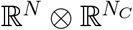.

The dynamics of the epidemic close to the disease-free state is captured by the infection propagator *P* (also an operator on 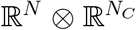), counting the possible transmission chains among hosts [27, 28, 30]. Following [28], we compute *P* as a function of *J* using the theory of nonautonomous linear systems on Eq. (2):

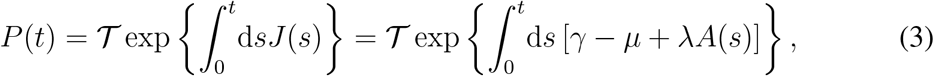

where 𝒯 is Dyson’s time-ordering operator (see Methods). The largest eigenvalue of *P* yields the epidemic threshold [27, 28, 30].

EGDs emerge as a graph-theoretical representation of the analytical treatment just presented. They are network representations that fully encode the infectious disease dynamics in real populations close to the critical spreading condition. They are composed by *N*_*C*_ nodes, each representing a disease compartment (except S), connected by single directed links (spontaneous transitions) or double directed links (transmission events) (Fig. 1b). Nodes can also have self-loops (single, for transitions to S; double, for transmissions entering the same node). EGDs completely bypass Eq. (1), as they can be built directly from the classic compartmental model and the network adjacency matrix with simple rules (steps 1-2 of Fig. 1d). Most importantly, computing the weighted adjacency matrix of the EGD (plus probability conservation) yields exactly the operator *J*(*t*) (Fig. 1c,d) appearing in the expression of the infection propagator of Eq. (3) (see Methods).

Under the simple representation of a weighted directed graph, EGDs hide a higherorder complexity. Unlike common graphs, link weights are not scalar but operators on ℝ^*N*^. Considering the relation between tensors and multilayer networks [42], we can interpret EGDs as graphs of layers: each node in the EGD (compartment) is a replica layer of the contact network, and links in the EGD are coupling operators among layers.

Operationally, EGDs make it possible to simplify different disease models and interventions, compare them, build equivalence classes. This is done through three diagram operations: `CUT`, `ZIP`, and `SHRINK`. They decompose diagrams into simpler parts, and compress complex disease progressions. Figure 2 describes in detail the diagram operations, which we present hereafter through specific epidemic applications. The corresponding proofs are reported in Methods. Finally – and more pragmatically – EGDs are a powerful tool for computing the epidemic threshold.

**Figure 2.**
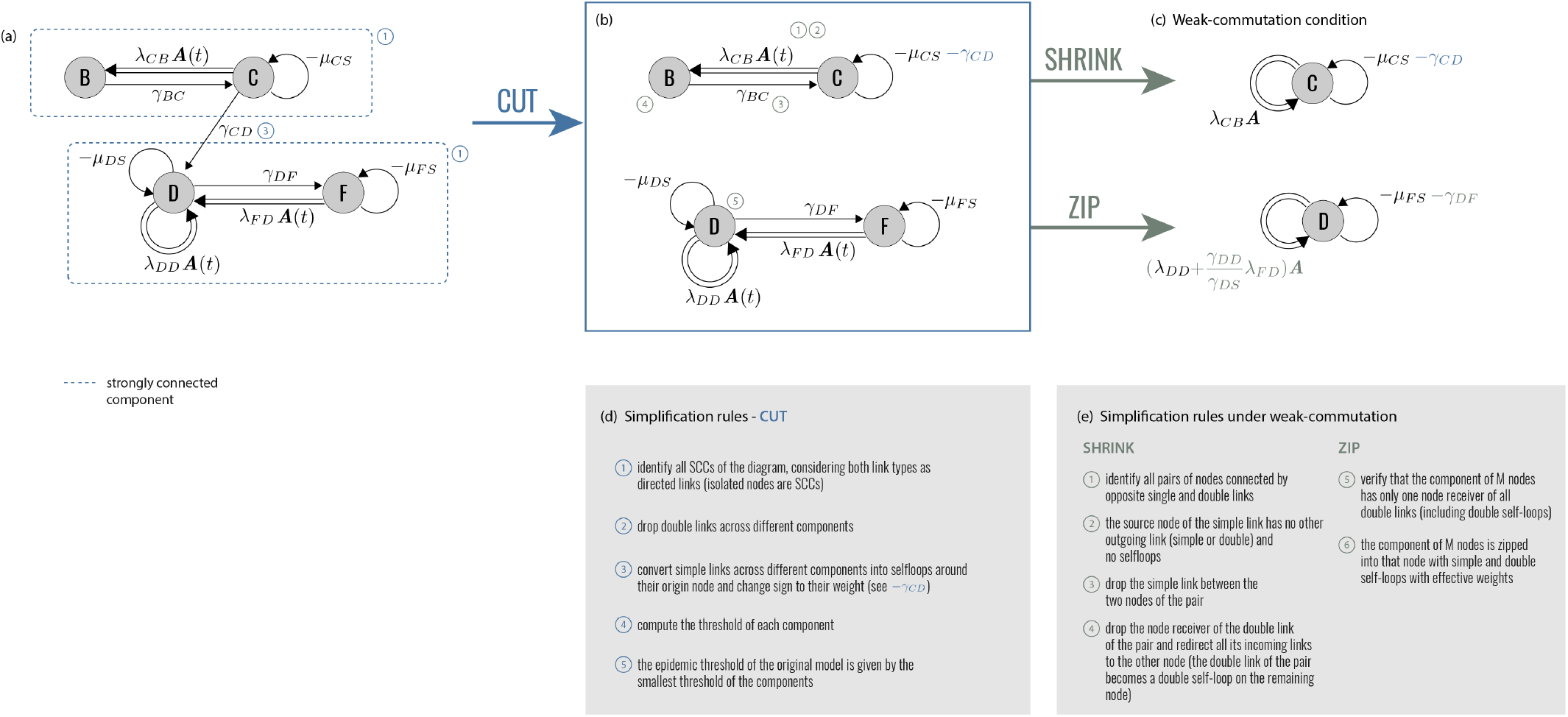
Diagram operations. **(a)** EGD of Fig. 1. The two strongly connected components are highlighted by blue rectangles. **(b)** Diagram operation `CUT` isolates the strongly connected components, which become disjoint subdiagrams. The epidemic threshold is then the smallest among the thresholds of each subdiagram. **(c)** Under the weak-commutation condition, each subdiagram can be further reduced through (i) diagram operation `SHRINK` (top) and (ii) diagram operation `ZIP` (bottom), leading to two EGDs of SIS models. Their transition rates are renormalized by the diagram operations. **(d), (e)** General rules of the three operations. Numbers refer to the steps to be performed for each operator; some of these steps are illustrated in panels a-c with the same numbers.

## Syphilis

As a first application, we consider syphilis spreading on a network of sexual contacts. Syphilis is a bacterial STI that is still widespread in resource-constrained countries and resurgent in high-income countries in specific risk groups [8]. After infection, the disease progresses through an incubation period lasting on average 3-4 weeks but with large fluctuations (10-90 days) [43]. It is then followed by two infective stages – primary and secondary syphilis. If untreated, syphilis enters latency, potentially leading to severe complications in the tertiary stage. Most models greatly simplify this disease progression and consider early stages only, under the assumption of antibiotic treatment, through susceptible-infectious-susceptible [44] or susceptible-infectious-recovered-susceptible approaches [45]. Incubation period is neglected, because complexity is shifted from disease progression towards capturing the heterogeneity of human sexual behavior [44].

Here we want to keep such complexity in *A*^*ij*^(*t*) while also preserving the progression of the early stages of syphilis infection. Introducing the incubation period following infection is often the first step into building a multi-stage disease history in realistic epidemic contexts [46]. Following the grammar of Fig. 1, we build the epidemic graph diagram of a susceptible-exposed-infectious-recovered-susceptible (SEIRS), where E corresponds to individuals incubating the disease but not yet infectious (Fig. 3). Diagram operation `CUT` allows the isolation of two strongly connected components, one containing only R, and one containing E and I (Fig. 3b). The first has no transmission terms. It is thus always below threshold, and can be dumped. The second leads to the simplified version of the Jacobian of the original diagram (Fig. 3c).

**Figure 3.**
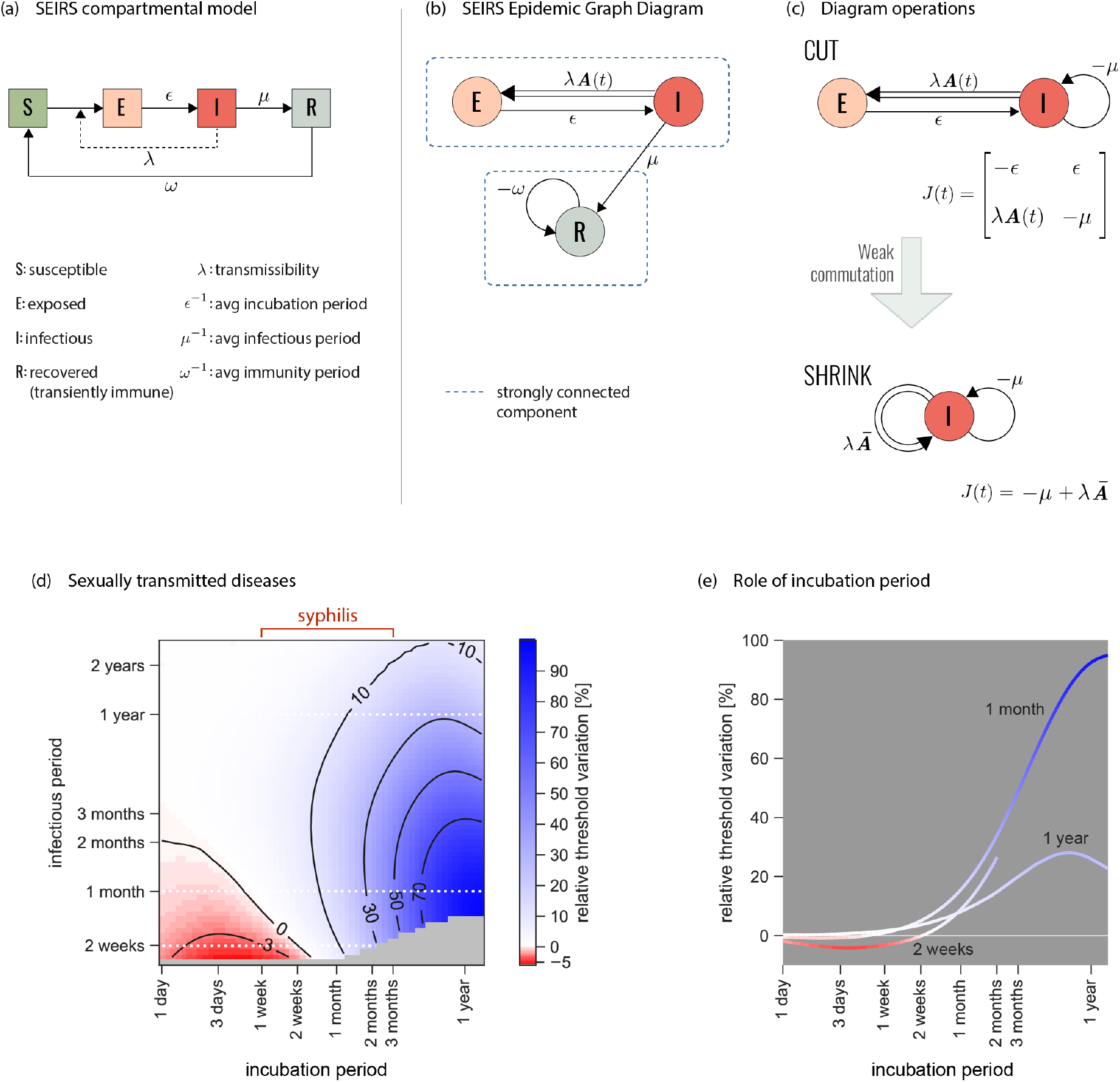
Epidemic threshold for syphilis. **(a)** Classic representation of the susceptibleexposed-infectious-recovered-susceptible (SEIRS) compartmental model here used to model the spread of a general STI. Parameterization for syphilis is provided in panel d. E represents the class of individuals exposed to the infection who are incubating the disease, before becoming infectious (I). R corresponds to temporary immunity. Rates are also shown. **(b)** Associated EGD. Strongly connected components are highlighted by blue rectangles. **(c)** Simplification of the EGD through diagram operations. The reduced diagram after `CUT` is shown with its associated Jacobian (top; the R component does not contribute to the threshold and is discarded). Under the weak-commutation condition, `SHRINK` further reduces the diagram to an EGD of an SIS model (bottom). **(d)** Relative difference in the prediction of the epidemic threshold in the full model (that is equivalent to the top diagram of panel c, after `CUT`) vs. the `CUT`+`SHRUNK` diagram (bottom diagram of panel c) obtained under the weak-commutation condition (i.e. removing the role of incubation). Results are obtained for an STI spreading on sexual network from real data [21], exploring different lengths of infectious period (i.e. time-to-treatment) and incubation period. Parameterization for syphilis infection is highlighted. A negative relative threshold variation indicates the threshold is lower in the full model compared to the weak-commutation one, i.e. the more realistic model predicts a higher risk than the approximated one. Both negative and positive variations are observed in the region of parameters corresponding to syphilis infection. The gray region of the plot indicates that the system is always below threshold. **(e)** Relative threshold variation as a function of the incubation period for the three infection durations (2 weeks, 1 month, 1 year) corresponding to the white dashed lines in panel d.

The diagram can however be further simplified if the network of contacts satisfies the weak-commutation condition [28] (see Methods). This applies to homogeneous mixing, static and annealed networks [28], and some temporally evolving network models such as the activity-driven model [47]. Under this condition, diagram operation `SHRINK` allows the removal of those nodes (compartments) having only one outgoing link in the EGD, as long as the link is single (Fig. 2c,e). Applied to syphilis, `SHRINK` eliminates E and reduces the original model to an SIS (Fig. 3c), whose threshold can be easily solved with the scalar version of the infection propagator approach [27, 28, 30]. Under weak-commutation, the incubation period therefore plays no role in the condition for endemic circulation, extending to temporal networks the result previously restricted to static and annealed networks [48]. For example, approaches neglecting syphilis incubation within the homogeneous mixing approximation [45] will not be biased if temporal correlations in sexual activity are negligible. If instead such correlations exist, errors are to be expected. To evaluate such errors, we consider syphilis circulating on a sexual network between Brazilian sex workers and clients [21] and measure the relative variation of the epidemic threshold computed on the `CUT` and on the `CUT`+`SHRUNK` diagrams of Fig. 3c. Within the variation of incubation duration reported in syphilis infected individuals [43], we find errors in elimination predictions ranging from −5% to 40% (Fig. 3d,e), if the model neglects incubation. The simple addition of one timescale in a non-infectious compartment induces an interplay with the timescales of the contacts and of the other disease stages, either boosting or hampering the conditions for syphilis spread. In a wave analogy, incubation acts in tuning the phase shift between disease and network, from in-phase (boosting) to counterphase (hampering) [49].

In the absence of treatment, or if treatment fails (e.g. for resistance emergence), later stages of syphilis infection need to be considered. The Supplementary Information reports the corresponding EGD and shows that even in that scenario the diagram can be simplified to the EGD of an SIS model (section S1).

The effect observed here for syphilis is relevant to other diseases, especially those for which natural history is poorly known, typically in an early phase of the outbreak. This is the case, for example, of the current monkeypox outbreak: estimates from endemic areas are not directly applicable to the ongoing epidemic, which features different spreading routes, symptomatology, and risk factors for acquisition [10]. In the absence of reliable estimates of disease time scales (incubation, generation time, detection) and stages (asymptomatic transmission), the analysis of present and future epidemic scenarios requires a flexible and agile theoretical framework, as the one developed here, to account for uncertainties.

## Pandemic influenza

Progression from latent infection to active disease may be far more complex than what illustrated for syphilis. Fully describing the natural history of influenza in human hosts, for example, requires a progression from exposed (E) to pre-symptomatic (P, infectious without showing symptoms yet), asymptomatic (A) or symptomatic (I) infectious, and recovered (R) (Fig. S3) [6]. Next to the inclusion of the incubation period, the distinction among infectious individuals is critical for public health interventions. Only symptomatic infectious (I) can be detected in the population and administered antivirals. P, A cannot. Antivirals treat severe forms of seasonal influenza, but most importantly represent the first line of defense against an emerging pandemic, as they mitigate morbidity and mortality at the population level [5, 6]. For this reason, neuraminidase inhibitors – the most common influenza antivirals – are stockpiled for pandemic preparedness [4]. However, large-scale antiviral administration may favor the emergence and spread of antiviral resistant influenza strains, compromising individual treatment and pandemic control [50]. Combined administration of two antivirals (combination therapy) has been proposed to defuse this threat, as opposed to monotherapy [6]. To study its effects in altering the pandemic potential of the circulating pandemic strain, we build the EGD of the full pandemic influenza disease progression model, adding antiviral combination therapy and emergence of drug resistance. Symptomatic infectious individuals (I) are treated with a certain probability *p*_*T*_. Mutation can occur in treated individuals giving rise to mono-resistant (to either drug, 1 or 2) and multi-resistant variants, with fitness cost *ϕ*. In the context of current pandemic preparedness [4], drugs 1 and 2 would correspond to oseltamivir Tamiflu and zanamivir Relenza. The full model is described in the Supplementary Information (section S2). It includes 25 compartments, 19 of which describe infectious stages.

Despite the model complexity, four strongly connected components can be identified in the corresponding EGD (Fig. 4a, neglecting the trivial component composed only of R, as before). They summarise the infection dynamics of the wild-type, the two monoresistant, and the multi-resistant strains. Moreover, the diagrams of the first three are isomorphic, underlying the equivalence of the associated dynamics. The `CUT` operation provides then the epidemic threshold as the lowest among the critical conditions of the different strains. Each condition can be computed from data on population structure, *A*(*t*), and with empirically-informed strain-specific parameter values (section S2).

**Figure 4.**
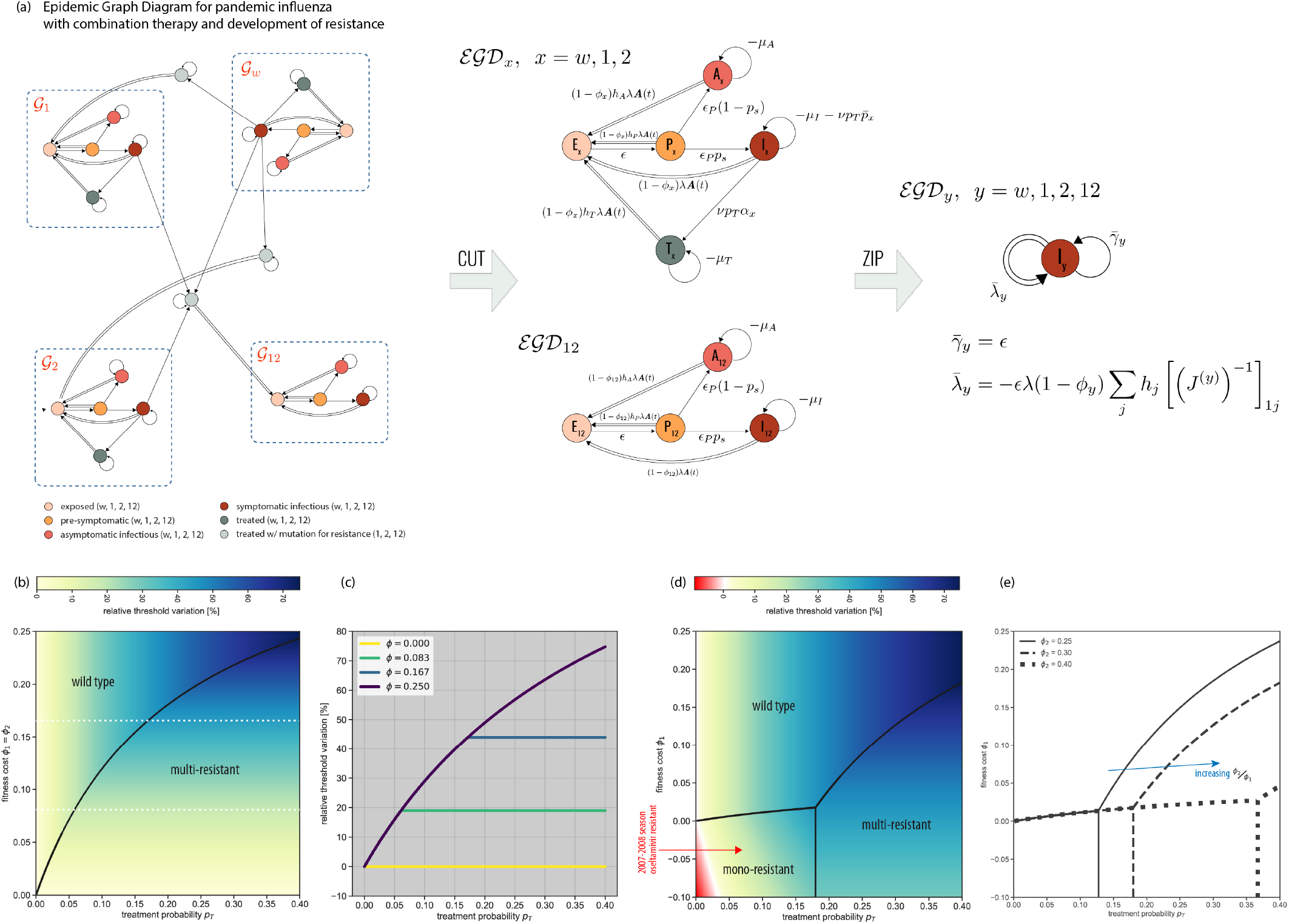
Epidemic threshold for pandemic influenza with antiviral combination therapy and emergence of resistance. **(a)** EGD of the pandemic influenza model and associated simplifications. The model includes four strains – wild-type, two mono-resistant, and one multi-resistant. The recovered (R) compartment has already been `CUT` out for the sake of visualization (see also Fig. 3b). The four strongly connected components (blue rectangles) correspond to the dynamics of each strain, independently. They can be isolated using `CUT`. Under the weak-commutation condition, the `ZIP` operation reduces each of the four subdiagrams to an SIS-like EGD with renormalized strain-specific parameters. Parameter definitions and values appear in Supplementary Tables S1-S3 and S5. **(b)** Relative threshold variation under treatment (*p*_*T*_ *>* 0) compared to no treatment (*p*_*T*_ = 0) as a function of *p*_*T*_ and fitness cost, assumed to be the same for both antivirals (*ϕ*_1_ = *ϕ*_2_). Positive relative threshold variation indicates the epidemic threshold is higher when antiviral drugs are used for therapy, i.e. the risk for a pandemic is reduced. The black line separates the two dominance regimes, for the wild-type strain and for the multi-resistant variant. **(c)** Relative threshold variation as a function of *p*_*T*_ along the values of fitness cost *ϕ*_1_ indicated by the white dashed lines in panel b. Treatment increases the epidemic threshold. But after a critical *p*_*T*_ the multi-resistant strain becomes dominant and further increasing treatment has no additional effect. **(d)** As panel b when fitness costs are specific to the drug, i.e. *ϕ*_2_ *> ϕ*_1_ [6]. A phase in which a mono-resistant strain dominates appears, differently from the situation depicted in panel b. In addition, when *ϕ*_1_ *<* 0 (resistance increases transmissibility), a region in parameter space emerges where threshold variation is negative (red region), i.e. the pandemic risk is increased by the use of antiviral drugs, due to the emergence of resistance. The red arrow indicates the parameter values estimated for the oseltamivir-resistant H1N1 strain, globally dominant in 2007-2008 [50]. **(e)** Boundaries of the three dominance phases of panel d when varying *ϕ*_2_. We report their analytical derivation in S2.2.

But the influenza diagram can be further simplified in most contexts. The short time scale of face-to-face proximity interactions along which influenza transmission can occur [22] makes annealed approximation, i.e. the weak-commutation assumption [28], the commonly adopted approximation. The diagram can then be fully compressed through the `ZIP` operation (Fig. 2c,e), because transmission events involve exclusively susceptible individuals entering the same compartment (here, exposed E). The difference between `ZIP` and `SHRINK` is that `ZIP` imposes global requirements on the diagram topology and compresses a full diagram, whereas `SHRINK` has only local requirements and merges two compartments at the time. The multi-resistant strain EGD and the three isomorphic EGDs (wild-type, and two mono-resistant strains), made of 4 and 5 compartments, respectively, are all `ZIP`ped into an EGD of an SIS model with renormalized transmission and recovery rates (Fig. 4a). The physical intuition is that multi-stage disease progression may change the speed of diffusion, but not whether the epidemic will break out or not, provided there are no dynamical interactions between the disease and the underlying contact network. Theoretically, we are able to disentangle the dynamics of the four different viral strains, analyze each of them separately, and show that they are equivalent (after appropriate parameter renormalization). Practically, we reduce a 25-compartment influenza model into a simple SIS model with substantial numerical gain: computing the largest eigenvalue of four *N* -dimensional matrices, instead of a 23*N* - dimensional matrix.

The factorization of the network component in the weak-commutation condition (section S2) allows us to make predictions on the dominant influenza variant by comparing the strain-specific thresholds against the scenario with no treatment (*p*_*T*_ = 0). Increasing treatment coverage helps controlling pandemic influenza, especially for high fitness costs of the resistant variants (Fig. 4b with *ϕ*_1_ = *ϕ*_2_). Above a certain value of *p*_*T*_, however, the multi-resistant strain dominates and the likelihood of its establishment is not affected anymore by the treatment coverage. The saturation effect depends on the fitness cost of the resistant strains (Fig. 4c).

Allowing fitness costs to depend on the specific drug (*ϕ*_1_ < *ϕ*_2_) leads to the emergence of an additional phase where the mono-resistant strain dominates, for small enough treatment probabilities and fitness costs (Fig. 4d). Notably, there exists a range of *ϕ*_1_ where increasing the intervention coverage leads to all possible phases – namely, either the wild-type dominates (small *p*_*T*_), or the mono-resistant strain (intermediate *p*_*T*_), or the multi-resistant variant (large *p*_*T*_). The dominance of a single mono-resistant strain is favoured by an increase in transmissibility following mutation (*ϕ*_1_ *<* 0), as expected. Though rare, such condition was observed in the 2007-2008 influenza season when an oseltamivir-resistant H1N1 variant emerged and rapidly spread, becoming the dominant H1N1 strain globally [50]. Dominance can happen also under reduced transmissibility of both mono-resistant strains (*ϕ*_1_ *>* 0), for a smaller region of parameter values. Large clusters of oseltamivir-resistant H1N1 variants were isolated in Japan in 2013-2014 influenza season that caused large community outbreaks but did not lead to large-scale dominance [50]. Increasing the difference between the two fitness costs magnifies the size of the mono-resistant phase (Fig. 4e).

The models presented so far are not age-structured. In the Supplementary Information (section S3) we use COVID-19 to show that EGDs can accommodate age-structured populations, a crucial feature for diseases for which exposure, transmission, or morbidity are age-dependent.

## Conclusions

The use of EGDs for pandemic influenza and syphilis show the versatility of the theoretical framework in solving the critical epidemic conditions while handling the full complexity of contact data, disease natural history, and interventions. EGDs share the same approximation of the infectious propagator approach, i.e. the quenched mean-field assumption [24, 26]. Its validity has been numerically tested for the critical condition [51], and specifically within the infection propagator framework [27]. All other results leading to the EGDs and their simplifications are obtained from the properties of the Jacobian, under no additional approximation. EGDs thus provide the analytics to predict public health risks at high granularity and to customise interventions, responding to the challenges of today’s public health.

## Data Availability

All data (parameter estimates) are reported in the manuscript and supplementary information.

## Acknowledgements

This study was partially supported by: the ANR projects COSCREEN (ANR-21-CO16-0005) and DATAREDUX (ANR-19-CE46-0008-03) to VC; EU Horizon 2020 grant MOOD (H2020-874850) to EV, CP, VC. We thank Alain Barrat and Mason Porter for useful discussions.

## Materials and Methods

### Infection propagator and epidemic threshold

We expand here the expression of the infection propagator of Eq. (3), which was derived using the theory of nonautonomous linear systems [52]. Dyson’s time-ordering operator [53] is defined as follows: 𝒯 *A*(*t*_1_)*A*(*t*_2_) = *θ*(*t*_1_−*t*_2_)*A*(*t*_1_)*A*(*t*_2_)+*θ*(*t*_2_−*t*_1_)*A*(*t*_2_)*A*(*t*_1_). *θ* is Heaviside’s step function. The time-ordered exponential of Eq. (3) is then a common representation of the following series:

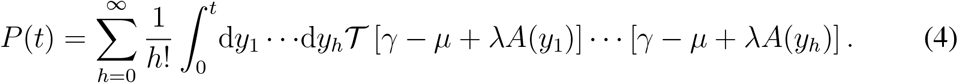

Time-ordering can be made explicit:

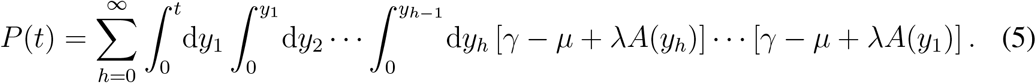

The infection propagator encodes the epidemic dynamics close to the epidemic threshold: 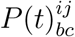 is the probability that *j* is in compartment *c* at time *t*, given that *i* is in compartment *b* at time *t* = 0, under the quenched mean-field assumption. We can now generalize what found in Ref. [27] for the SIS model, and compute the epidemic threshold. The epidemic threshold is the critical parameter surface separating the disease-free state to the endemic phase. In the SIS model it is a one-dimensional curve relating the transmission rate from the recovery rate. Here instead it is a (*N*_*C*_ − 1)-dimensional surface defined as follows:

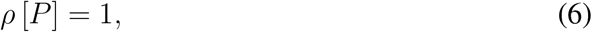

where *ρ* indicates the spectral radius, i.e., the largest eigenvalue. In practice, it is still possible to obtain an equation for one single parameter if all transmission rates *λ*_*bc*_ are expressed as functions of the baseline transmission rate, whose critical value gives the epidemic threshold. This is the case of the pandemic influenza model, for example, where transmissibility of each infectious compartment (e.g. P, A, treated symptomatic infectious individuals, mono-resistant or multi-resistant strains, etc.) is defined as a rescaling of the transmissibility of the symptomatic infectious individual (e.g. transmissibility of the mono-resistant strain is equal to the transmissibility of the wild-type, rescaled for the fitness cost, see Supplementary Information section S2).

To numerically compute the spectrum of *P*, an operator on 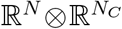, we interpret the tensor products in *J* as Kronecker products. The resulting matrix has dimension *NN*_*C*_, and has a block structure, with each block of dimension *N*, as shown in Fig. 1c. Formally, we are exploiting the isomorphism 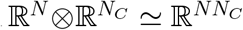, and this is equivalent to the supra-adjacency representation of multilayer networks [42]. When *γ* = 0, and *N*_*C*_ = 1 (scalar), we recover the infection propagator of the susceptible-infectious-susceptible model [28]. If the network obeys the *weak-commutation condition* [28, 54], both the infection propagator (Eq. (3)) and the threshold (Eq. (6)) simplify. The weak-commutation condition is defined in Ref. [28] as

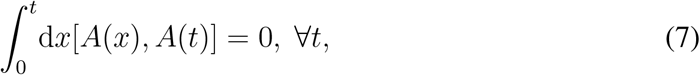

where [*A*(*x*), *A*(*t*)] = *A*(*x*)*A*(*t*) − *A*(*t*)*A*(*x*) is the standard matrix commutator. As Ref. [28] proves, this is true in two cases: i) temporal correlations in the evolution of *A*(*t*) are absent (network annealing); ii) the timescale of temporal correlations in the evolution of *A*(*t*) is much shorter than the timescale of the spread of the disease (timescale separation). In both cases it is possible to replace *A*(*t*) with the average adjacency matrix *Ā*, and sum the series in Eq. (3). The infection propagator is then the exponential of the Jacobian:

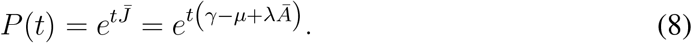

Its spectrum is the exponentiation of the spectrum of *J*. From this, the equation of epidemic threshold becomes

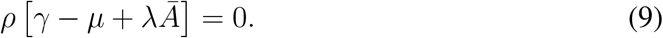

In case of an SIS model (*γ* = 0; *µ, λ* scalars), Eq. (9) becomes the well-known formula *λ* = *µ/ρ*[*Ā*] [24, 26].

Lastly, we comment on the term *f* (*x*) in Eq. (1). As stated in the paper, this does not contribute to the epidemic threshold because its contribution to the Jacobian in the disease-free state vanishes. In the classical representation of a compartmental model showed in Fig. 1a, type 3 transitions (i.e. transmissions infecting other compartments than S) would be contained in *f*. They would correspond to quadratic terms of the form *x*_*b*_*x*_*c*_. Since *f* can be ignored, this means that type 3 transitions can be ignored too, and never appear in EGDs.

### From the infection propagator to the EGD

We stated that epidemic graph diagram is a graphical representation of *J*, which can be obtained from the EGD by computing its weighted adjacency matrix (plus conservation condition), as shown in Fig. 1d:

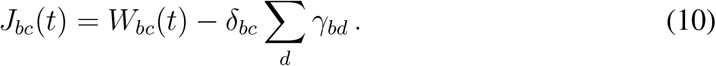

To proof the equivalence between the EGD and the infection propagator approach to analytically compute the epidemic threshold, we need to demonstrate that the Jacobian of the above equation is exactly *J*(*t*) = *γ* − *µ* + *λA*(*t*), i.e. the expression of the Jacobian obtained from the dynamical equations in the main paper. We note that the parameters *γ*_*bc*_ appearing in the general compartmental model (see Fig. 1a) are never diagonal: *b* ≠ *c*. Therefore, the weighted adjacency matrix of the EGD that appears in Eq. (10) is

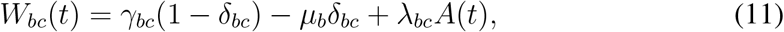

where we made explicit the off-diagonal nature of *γ*_*bc*_. The diagonal elements of operator *γ* in Eq. (3) are not parameters of the model, they are fixed by probability conservation, i.e. *γ*_*bb*_ = − ∑_*d*≠*b*_ *γ*_*bd*_. We therefore need to add this last term to the weighted adjacency matrix of the EGD to account for the diagonal entries of operator *γ* of Eq. (3), and this explains the nature of the second term in Eq. (10). This proves that the Jacobian built from the EGD is exactly the operator *J*(*t*) of the infection propagator.

### Diagram operations

We prove here the diagram operations described in the paper, and in Fig. 2. They are `CUT`, `ZIP`, and `SHRINK`. `CUT` decomposes EGDs in smaller subdiagrams using network approaches and allows the computation of the threshold on each subdiagram separately. `ZIP` compresses complex diseases progressions into SIS-like diagrams. `SHRINK` merges pair of nodes. `ZIP` exploits and transforms the global topology of the diagram, while `SHRINK` acts locally on pairs of compartments.

#### Proof of CUT

An EGD is a directed graph, and it is possible to order its nodes so that its adjacency matrix is block-upper-triangular, with the blocks representing the strongly connected components. *J* inherits the same property, if intended as the supra-adjacency matrix [42, 55] of the multilayer structure represented by the EGD, i.e., the adjacency matrix of the flattened graph exploiting 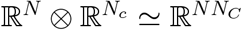. And so does *P*, because it is a convolution of *J*(*t*) at different times. It follows that it is possible to compute the spectrum of *P* as the union of the spectra of the blocks in the main diagonal, which are the subdiagrams obtained via `CUT`.

#### Proof of ZIP

We use the notation ℰ𝒢𝒟 for the mathematical representation of the diagram. Let us assume that weak-commutation condition holds (*A*(*t*) ≡ *Ā*), and the epidemic graph diagram ℰ𝒢𝒟 is made of a node *D* and a subdiagram ℰ𝒢𝒟_*X*_. Moreover, let us assume that ℰ𝒢𝒟_*X*_ contains only single links (its Jacobian is *γ*_*X*_ − *µ*_*X*_). Single links may exist from *D*, to any node in ℰ𝒢𝒟s_*X*_: We call 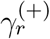 the weight of the single link from *D* to the *r*-th node of ℰ𝒢𝒟_*X*_. Single links may also exist from any node in ℰ𝒢𝒟_*X*_ to *D*: We call 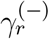 the weight of the single link from the *r*-th node of ℰ𝒢𝒟_*X*_ to *D*. The index *r* runs on the *N*_*c*_ −1 nodes of ℰ𝒢𝒟_*X*_. *D* may have both single and double self loops, whose weights we call −*µ*_0_ and *λ*_0_*Ā*, respectively. Double links may exist from any node in ℰ𝒢𝒟_*X*_ to *D*(*λ*_*r*_). The resulting *J* of the full EGD is

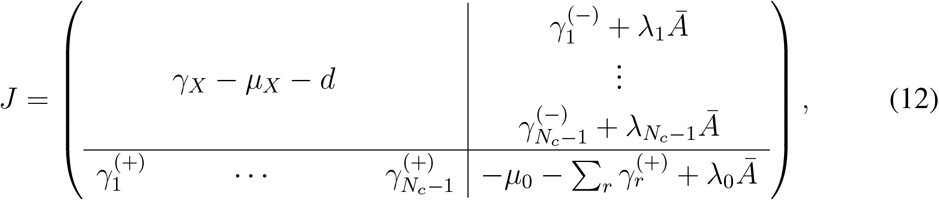

with 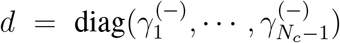. The threshold condition in timescale separation is det *J* = 0. Using block matrix determinant rules, this simplifies to a determinant in ℝ^*N*^ :

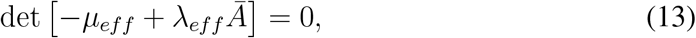

with

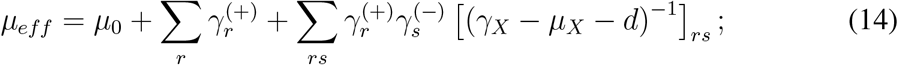

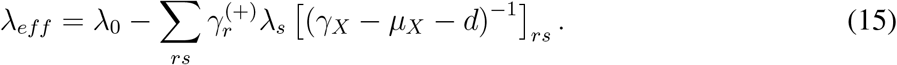

*γ*_*X*_ − *µ*_*X*_ − *d* is always invertible if we assume that the model is below the epidemic threshold in the absence of transmission. If it were not the case, the epidemic would be a trivial spontaneous generation of infected individuals. Or, in epidemiological terms, there would be only primary disease introductions. This means that *J*(*λ*_0_ = 0, *λ*_*r*_ = 0) is negative definite. By virtue of Sylvester’s criterion, this in turn implies that *γ*_*X*_ −*µ*_*X*_ −*d* is also negative definite, because all the leading principal minors of the latter are also leading principal minors of *J*. And since *γ*_*X*_ − *µ*_*X*_ does not depend on transmission, *γ*_*X*_ − *µ*_*X*_ − *d* is always negative definite, and so invertible. We complete the proof of `ZIP` by noting that Eq. (13) is equivalent to an SIS model with renormalized recovery and transmission rates *µ*_*eff*_, *λ*_*eff*_.

#### Proof of SHRINK

The proof is conceptually similar to ZIP’s. Let us assume that the epidemic graph diagram is composed of compartment *B* and a subgraph ℰ𝒢𝒟_*X*_, with Jacobian *γ*_*X*_ − *µ*_*X*_ + *λ*_*X*_*Ā*. Morever, let us assume that a single link *γ*_0_ goes from *B* to a node in ℰ𝒢𝒟_*X*_, which we label as the first (*r* = 1). *B* may have no other outgoing link, and no self loops. Any node in ℰ𝒢𝒟_*X*_ may have single and double links going to *B*. The Jacobian is

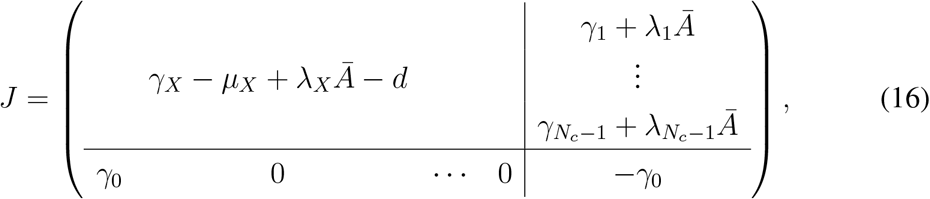

with 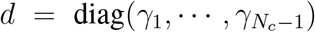. Using block matrix determinant rules, the condition det *J* = 0 is equivalent to the following *N*_*c*_ − 1 dimensional determinant

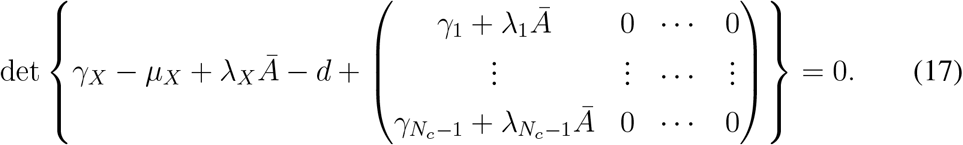

This is equivalent to a diagram in which *B* disappears, its outgoing link *γ*_0_ disappears, and all incoming links of *B* get rerouted onto the ancient target of link *γ*_0_.

## Notes

### Competing Interest Statement

The authors have declared no competing interest.

