## Supplementary Information for "Epidemic graph diagrams as analytics for epidemic control in the data-rich era"

[1]INSERM, Sorbonne Université, Institut Pierre Louis d'Epidémiologie et de Santé Publique, F75012, Paris, France.
[2]aizoOn Technology Consulting, 10146 Turin, Italy.

### 1 Syphilis

We illustrate here the compartmental model of syphilis introduced in [1] considering the full disease progression. Compared to the model we use in the main paper, it differentiates between primary and secondary stages, and includes later stages of the disease, such as latent syphilis, reactivation of secondary syphilis, tertiary syphilis. The classic representation of the compartmental model is shown in Figure S1. Once infected, susceptible individuals enter the exposed compartment (E) before developing primary syphilis infection (compartment $I_1$). If not treated, they then progress to secondary syphilis ($I_2$). After that, a long period of spontaneous remission follows (latent syphilis L). The last stage of untreated syphilis is then tertiary syphilis ($I_3$). Before ending in $I_3$ some individuals present recurrent episodes of secondary syphilis ($I_R$). The latent stage preceding $I_R$ is in general shorter and is modelled with a different compartment (branch between L and $L_R$) in the figure.

Treatment may occur at any time during the disease progression. Upon treatment, individuals with primary and secondary infection are cured and become susceptible again, while individuals in compartments $L_R$, L, $I_R$ and $I_3$ retain temporary immunity after cure. Transition parameters are indicated in Figure S1.

We assume that individuals with primary and secondary infection ($I_1$, $I_2$ and $I_R$) have the same transmission rate $\lambda$, and latent and tertiary syphilis are not infectious. However, the EGD could be easily modified to accommodate violations of these assumptions, which would simply change the value of the final effective parameters in Eqs. (4) and (5) here below.

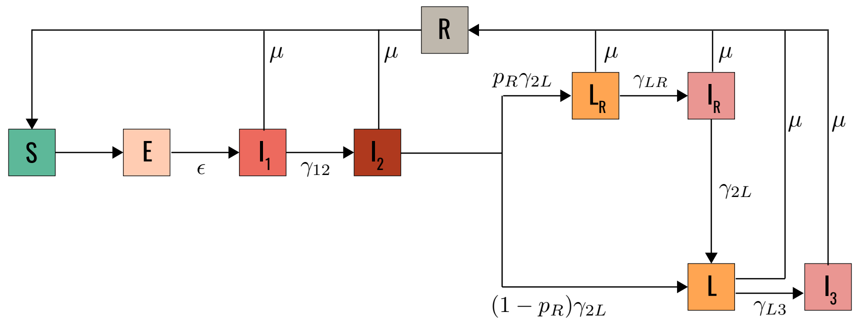


Figure S1: Compartmental model of syphilis considering the full natural history of the disease [1].

The EGD of the full compartmental model of syphilis is shown in Figure S2. We first `CUT` the diagram as shown by the two strongly connected components highlighted in the figure. Compartments R, L, and $I_3$ can be CUT out leaving a remaining subdiagram of 5-nodes (top component, nodes: $E, I_1, I_2, L_R, I_R$). If the weak-commutation condition holds, we ZIP this subdiagram and obtain an EGD of an SIS model whose effective parameters (see Methods of main paper) are

$$\mu_{eff} = \epsilon; \tag{1}$$

$$\lambda_{eff} = -\epsilon \sum_s \lambda_s (J_X^{-1})_{1s}, \tag{2}$$

where $J_X$, the Jacobian of the subdiagrams generated by nodes $I_1, I_2, L_R, I_R$, is given by

$$J_X = \begin{pmatrix} -\mu - \gamma_{12} & \gamma_{12} & 0 & 0 \\ 0 & -\mu - \gamma_{2L} & p_R\gamma_{2L} & 0 \\ 0 & 0 & -\mu - \gamma_{LR} & \gamma_{LR} \\ 0 & 0 & 0 & -\mu - \gamma_{2L} \end{pmatrix}. \tag{3}$$

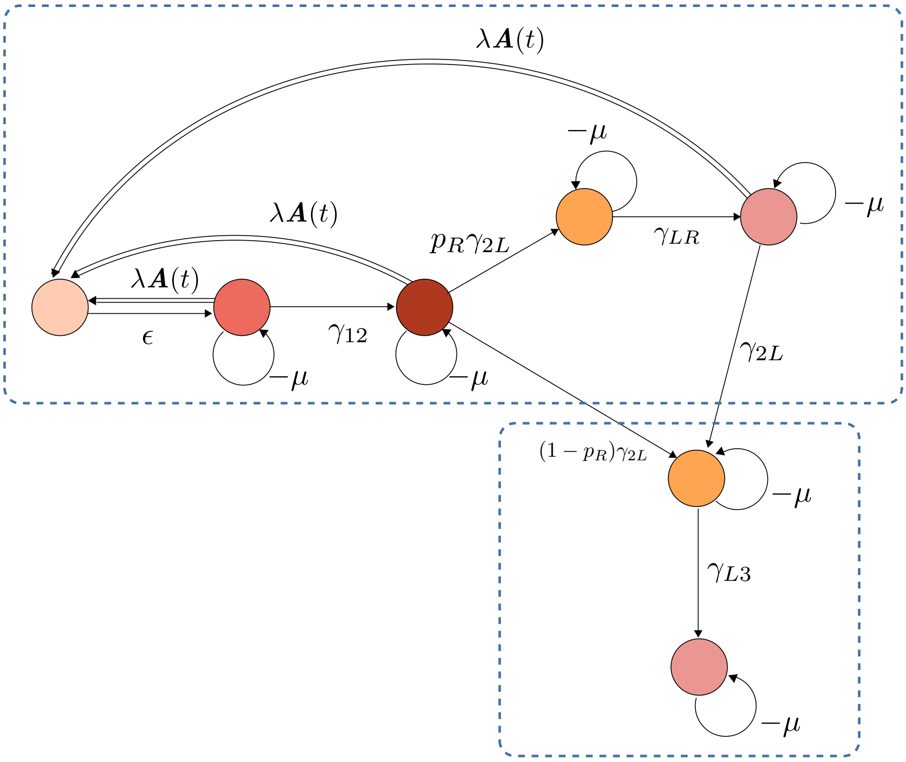


Figure S2: EGD of the syphilis model of Fig. S1.

It is convenient to use the epidemic threshold formula of the SIS model under weak-commutation condition to simplify the parameters further: $\lambda_{eff}/\mu_{eff}$ =

$1/\rho[\bar{A}]$. From this we can simplify both $\lambda_{eff}$ to $\mu_{eff}$, and remove $\epsilon$: under weak-commutation the incubation period does not impact the threshold, as we showed in the main paper for the simpler SEIRS model. Using the scaling $\lambda_{eff}/\mu_{eff} = 1/\rho[\bar{A}]$, we can also redefine the effective parameters so that $\lambda_{eff}$ matches syphilis transmissibility $\lambda$, and $\mu_{eff}$ contains the multistage corrections:

$$\lambda_{eff} = \lambda; \tag{4}$$

$$\mu_{eff} = (\mu + \gamma_{12}) \left\{ 1 + \frac{\gamma_{12} \left[ \mu(\mu + \gamma_{LR}) + \gamma_{2L}(\mu + (1 + p_R)\gamma_{LR}) \right]}{(\mu + \gamma_{LR})(\mu + \gamma_{2L})^2} \right\}^{-1} \tag{5}$$

### 2 Influenza

We model influenza and antiviral treatment following the approach of Ref. [2] (Figure S3). We consider two antiviral drugs and four strains with different resistance profiles:

- wild-type (subscript $w$);
- mono-resistant: resistant to either drug (subscript 1 or 2);
- multi-resistant: resistant to both drugs (subscript 12).

Susceptible individuals (S) can get infected by one strain and enter the exposed compartment ($\mathrm{E_y}$ with $\mathrm{y} = \mathrm{w}, 1, 2, 12$). After an average latency period $\epsilon^{-1}$ individuals become pre-symptomatic ($\mathrm{P_y}$) – i.e. infectious, with no clinical manifestation. A proportion $p_S$ of them develops symptoms with rate $\epsilon_P$ and enters the $\mathrm{I_y}$ compartment, while the rest remains asymptomatic ($\mathrm{A_y}$ compartment). Asymptomatic individuals have a shorter infectious duration compared to symptomatic individuals ($\mu_A^{-1} < \mu_I^{-1}$).

Upon onset of symptoms, treatment is administered with rate $\nu$ to a fraction of symptomatic, selected with probability $p_T$. Treated individuals who do not develop resistance (additional resistance if resistance to one drug is already present) go from $\mathrm{I_y}$ to $\mathrm{T_y}$. Those who do go from $\mathrm{I_y}$ to the treated infectious compartment corresponding to a different strain. As an example, the following outcome is possible for $\mathrm{I_w}$ individuals:

- they enter the compartment $\mathrm{T_w}$ (treated, infected with the wilde-type virus) with rate $\nu p_T(1-p_1)(1-p_2)$, where $p_1$ ($p_2$) is the probability of developing resistance to drug 1 (2);
- they enter the compartment $\mathrm{T_1^m}$ (or $\mathrm{T_2^m}$) with rate $\nu p_\mathrm{T} p_1(1-p_2)$ ($\nu p_\mathrm{T} p_2(1-p_1)$) corresponding to treated individuals infected with mutant virus of type 1 (2), if resistance is developed upon treatment;

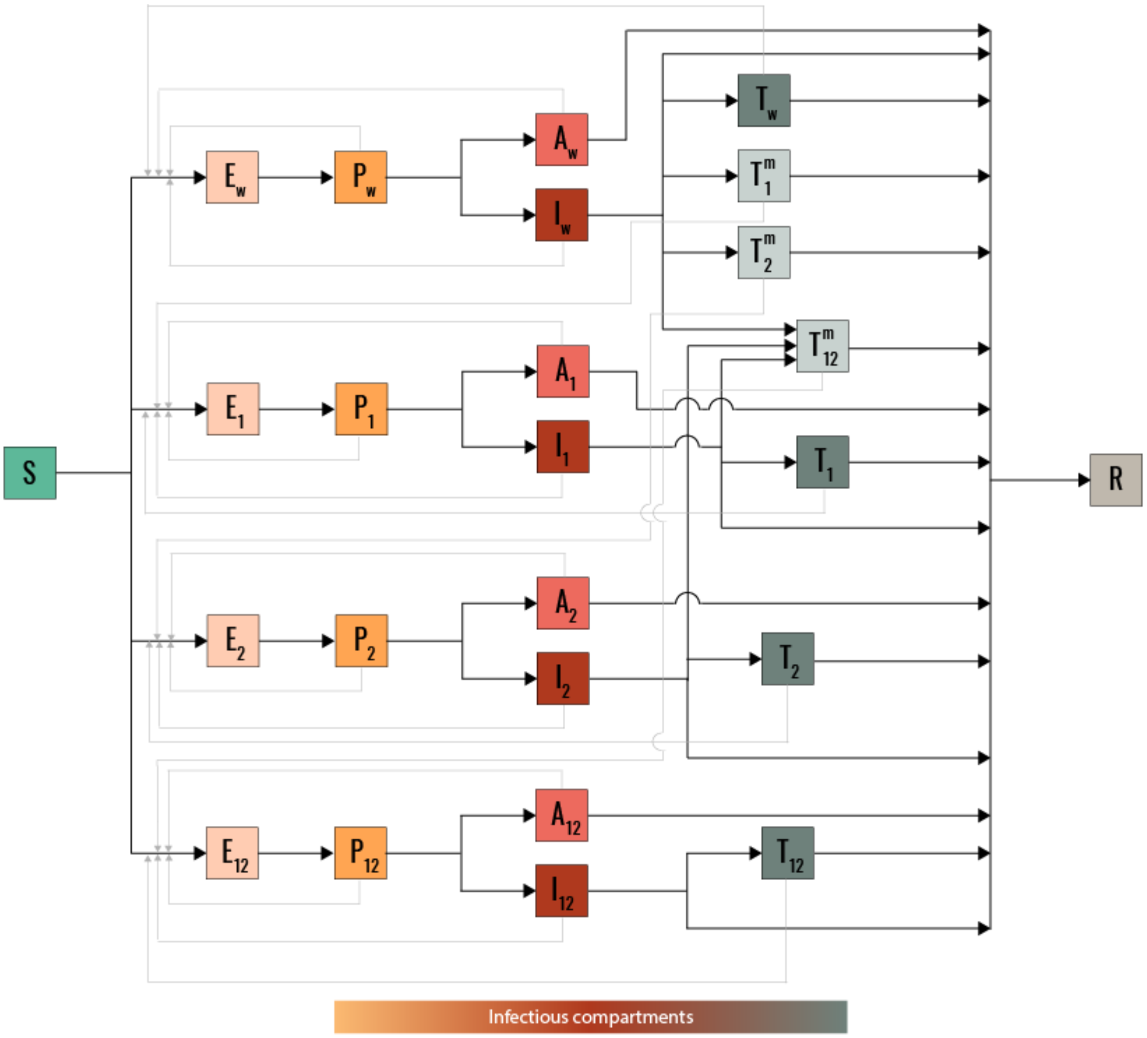


Figure S3: Compartmental model for pandemic influenza with antiviral combination therapy and development of drug resistance [2].

- they enter the compartment $\mathrm{T}_{12}^{\mathrm{m}}$ with rate $\nu p_{\mathrm{T}} p_1 p_2$ corresponding to treated individuals infected with the multi-resistant strain, if multi-resistance is acquired;

- they go untreated, i.e. transit directly from $\mathrm{I_w}$ to the recovered R compartment with rate $\mu_I(1-p_T)$, where $\mu_I$ is the recovery rate of untreated infectious individuals.

Mutations from mono-resistant to the wild-type or from multi-resistant to mono-resistant are not considered in the model [2]. We assume treatment to reduce duration of infection, with $\tau$ being the difference between the overall infection duration in treated and untreated individuals, i.e. $\mu_I^{-1} = \nu^{-1} + \mu_T^{-1} + \tau$. Treatment is still effective for mono-resistant strains since we assume a combined therapy. However, it has no effect on multi-resistant strain. Thus the overall infectious duration for the latter is the same as untreated individuals, i.e. $\mu_I^{-1} = \nu^{-1} + \mu_{mm}^{-1}$.

Transmissibility depends on strain and degree of symptoms. Being $\lambda$ the transmission rate of $\mathrm{I_w}$ taken as baseline value, transmission in the pre-symptomatic, asymptomatic and treated class is rescaled of a factor $h_P$, $h_A$ and $h_T$, respectively. At the same time, mutation may alter the transmission potential of the virus. Mutant strains are often (but not always) less fit [2, 3], thus we introduce the fitness cost parameter $\phi_1$ ($\phi_2$), and we explore both positive and negative values, where negative fitness cost means increased transmissibility upon mutation.

Parameters and their values are summarised in Table S1. For the analysis of Fig. 4b-e of the main paper we parametrise the model as in [2, 4, 5] and vary the values of $\phi_1$, $\phi_2$ and $p_T$ computing the corresponding threshold value of $\lambda$.

#### 2.1 `CUT` of pandemic influenza EGD

At first, `CUT` separates the recovered compartment from the remaining part of the $\mathcal{EGD}$ – see Fig. 4a of main paper. The latter can be then further cut into 4 diagrams: $\mathcal{EGD}_w, \mathcal{EGD}_1, \mathcal{EGD}_2, \mathcal{EGD}_{12}$. The weights of the links are reported in Table S2 ($\mathcal{EGD}_w, \mathcal{EGD}_1, \mathcal{EGD}_2$), and Table S3 ($\mathcal{EGD}_{12}$).

#### 2.2 `ZIP` of pandemic influenza EGD

All four submodels of the influenza model are `ZIP`pable. The effective parameters $\mu_{eff}, \lambda_{eff}$ of $\mathcal{EGD}_w, \mathcal{EGD}_1, \mathcal{EGD}_2, \mathcal{EGD}_{12}$ are

$$\mu_{eff}^{(x)} = \epsilon; \tag{6}$$

$$\lambda_{eff}^{(x)} = -\epsilon\lambda(1-\phi_x)\sum_j h_j \left[\left(J^{(x)}\right)^{-1}\right]_{1j}, \tag{7}$$

| name | type | description | value/notes |
|---|---|---|---|
| | | ***transmission*** | |
| $\lambda$ | transmission rate | $=\lambda(\mathrm{I_w})$, baseline | threshold value computed |
| $h_P$ | correction for pre-sym | $=\lambda(\mathrm{P_x})/\lambda(\mathrm{I_x})$ | $0.68$ |
| $h_A$ | correction for asym | $=\lambda(\mathrm{A_x})/\lambda(\mathrm{I_x})$ | $0.36$ |
| $h_T$ | correction for treated | $=\lambda(\mathrm{T_x})/\lambda(\mathrm{I_x})$ | $0.67$ |
| $\phi_1$ | fitness cost | $=1-\lambda(\mathrm{I_1})/\lambda(\mathrm{I_w})$ | explored |
| $\phi_2$ | fitness cost | $=1-\lambda(\mathrm{I_2})/\lambda(\mathrm{I_w})$ | explored |
| | | ***branching*** | |
| $p_S$ | prob symptoms | | $0.67$ |
| $p_T$ | prob treatment | | explored |
| $p_1$ | prob resistance to 1 | | $10^{-2}, 10^{-1}$ |
| $p_2$ | prob resistance to 2 | | $10^{-2}, 10^{-1}$ |
| $\tau$ | less inf. per. due to treatment | | $1\,day$ |
| | | ***spontaneous transitions*** | |
| $\epsilon$ | transition rate | E to P | $(0.98\,day)^{-1}$ |
| $\epsilon_P$ | transition rate | P to A, I | $(0.5\,day)^{-1}$ |
| $\nu$ | treatment rate | | $(0.5\,day)^{-1}$ |
| $\mu_I$ | recovery rate | for I | $(2.9\,day)^{-1}$ |
| $\mu_A$ | recovery rate | for A | $(0.5\,day)^{-1}$ |
| | | ***derived variables*** | |
| $\mu_T$ | recovery rate | for T | from $\mu_I^{-1}=\nu^{-1}+\mu_T^{-1}+\tau$ |
| $\mu_m$ | recovery rate | for $\mathrm{T_1^{(m)}}, \mathrm{T_2^{(m)}}$ | $=\mu_T$ |
| $\mu_{mm}$ | recovery rate | for $\mathrm{T_{12}^{(m)}}$ | from $\mu_I^{-1}=\nu^{-1}+\mu_{mm}^{-1}$ |

Table S1: Parameters of the pandemic influenza model. $\lambda(C)$ is the transmissibility of compartment $C$. Subscript $x$ takes values $w, 1, 2$. Parameter values from Ref. [2, 4, 5]

| source | target | type | value |
|---|---|---|---|
| | | $\mathcal{EGD}_w$ | |
| E | P | s | $\epsilon$ |
| P | A | s | $\epsilon_P(1-p_S)$ |
| P | I | s | $\epsilon_P p_S$ |
| I | T | s | $\nu p_T(1-p_1)(1-p_2)$ |
| A | A | s | $-\mu_A$ |
| I | I | s | $-\mu_I - \nu p_T[1-(1-p_1)(1-p_2)]$ |
| T | T | s | $-\mu_T$ |
| P | E | d | $h_P \lambda A(t)$ |
| I | E | d | $\lambda A(t)$ |
| A | E | d | $h_A \lambda A(t)$ |
| T | E | d | $h_T \lambda A(t)$ |
| | | $\mathcal{EGD}_1$ | |
| E | P | s | $\epsilon$ |
| P | A | s | $\epsilon_P(1-p_S)$ |
| P | I | s | $\epsilon_P p_S$ |
| I | T | s | $\nu p_T(1-p_2)$ |
| A | A | s | $-\mu_A$ |
| I | I | s | $-\mu_I - \nu p_T p_2$ |
| T | T | s | $-\mu_T$ |
| P | E | d | $(1-\phi_1)h_P \lambda A(t)$ |
| I | E | d | $(1-\phi_1)\lambda A(t)$ |
| A | E | d | $(1-\phi_1)h_A \lambda A(t)$ |
| T | E | d | $(1-\phi_1)h_T \lambda A(t)$ |
| | | $\mathcal{EGD}_2$ | |
| E | P | s | $\epsilon$ |
| P | A | s | $\epsilon_P(1-p_S)$ |
| P | I | s | $\epsilon_P p_S$ |
| I | T | s | $\nu p_T(1-p_1)$ |
| A | A | s | $-\mu_A$ |
| I | I | s | $-\mu_I - \nu p_T p_1$ |
| T | T | s | $-\mu_T$ |
| P | E | d | $(1-\phi_2)h_P \lambda A(t)$ |
| I | E | d | $(1-\phi_2)\lambda A(t)$ |
| A | E | d | $(1-\phi_2)h_A \lambda A(t)$ |
| T | E | d | $(1-\phi_2)h_T \lambda A(t)$ |

Table S2: Links of the subdiagrams $\mathcal{EGD}_w, \mathcal{EGD}_1, \mathcal{EGD}_2$, resulting from the `CUT`. Parameters are described in Table S1.

| source | target | type | value |
|---|---|---|---|
| | | $\mathcal{EGD}_{12}$ | |
| E | P | s | $\epsilon$ |
| P | A | s | $\epsilon_P(1-p_S)$ |
| P | I | s | $\epsilon_P p_S$ |
| A | A | s | $-\mu_A$ |
| I | I | s | $-\mu_I$ |
| P | E | d | $(1-\phi_1)(1-\phi_2)h_P\lambda A(t)$ |
| I | E | d | $(1-\phi_1)(1-\phi_2)\lambda A(t)$ |
| A | E | d | $(1-\phi_1)(1-\phi_2)h_A\lambda A(t)$ |

Table S3: Links of the subdiagram $\mathcal{EGD}_{12}$, resulting from the CUT. Parameters are described in Table S1.

where $x$ takes values $w, 1, 2, 12$; $J^{(x)}$ is the Jacobian of $\mathcal{EGD}_x - \{E\}$, $\phi_x$ is the fitness cost corresponding to $\mathcal{G}_x$ and $h = (h_P, h_A, 1, h_T)$.

These are the relevant Jacobian matrices:

$$J_w = \begin{pmatrix} -\epsilon_P & \epsilon_P(1-p_S) & \epsilon_P p_S & 0 \\ 0 & -\mu_A & 0 & 0 \\ 0 & 0 & -\mu_I - \nu p_T & \nu p_T(1-p_1)(1-p_2) \\ 0 & 0 & 0 & -\mu_T \end{pmatrix} \tag{8}$$

$$J_1 = \begin{pmatrix} -\epsilon_P & \epsilon_P(1-p_S) & \epsilon_P p_S & 0 \\ 0 & -\mu_A & 0 & 0 \\ 0 & 0 & -\mu_I - \nu p_T & \nu p_T(1-p_2) \\ 0 & 0 & 0 & -\mu_T \end{pmatrix} \tag{9}$$

$$J_2 = \begin{pmatrix} -\epsilon_P & \epsilon_P(1-p_S) & \epsilon_P p_S & 0 \\ 0 & -\mu_A & 0 & 0 \\ 0 & 0 & -\mu_I - \nu p_T & \nu p_T(1-p_1) \\ 0 & 0 & 0 & -\mu_T \end{pmatrix} \tag{10}$$

$$J_{12} = \begin{pmatrix} -\epsilon_P & \epsilon_P(1-p_S) & \epsilon_P p_S \\ 0 & -\mu_A & 0 \\ 0 & 0 & -\mu_I \end{pmatrix}. \tag{11}$$

And the first row of their inverses is

$$\left[\left(J^{(w)}\right)^{-1}\right]_{1j} = -\left(\epsilon_P^{-1}, (1-p_S)\mu_A^{-1}, p_S\left(\mu_I + \nu p_T\right)^{-1}, p_S p_T \nu (1-p_1)(1-p_2)\left(\mu_I + \nu p_T\right)^{-1}\mu_T^{-1}\right);$$

$$\left[\left(J^{(1)}\right)^{-1}\right]_{1j} = -\left(\epsilon_P^{-1}, (1-p_S)\mu_A^{-1}, p_S\left(\mu_I + \nu p_T\right)^{-1}, p_S p_T \nu (1-p_2)\left(\mu_I + \nu p_T\right)^{-1}\mu_T^{-1}\right);$$

$$\left[\left(J^{(2)}\right)^{-1}\right]_{1j} = -\left(\epsilon_P^{-1}, (1-p_S)\mu_A^{-1}, p_S\left(\mu_I + \nu p_T\right)^{-1}, p_S p_T \nu (1-p_1)\left(\mu_I + \nu p_T\right)^{-1}\mu_T^{-1}\right);$$

$$\left[\left(J^{(12)}\right)^{-1}\right]_{1j} = -\left(\epsilon_P^{-1}, (1-p_S)\mu_A^{-1}, p_S\mu_I^{-1}, 0\right). \tag{12}$$

The vector $\left[\left(J^{(12)}\right)^{-1}\right]_{1j}$ is actually 3-dimensional. We have added a zero fourth entry to so that the general scalar product in Eq. (7) holds.

$$\omega = h_P/\epsilon_P + (1-p_S)h_A/\mu_A, \tag{13}$$

$$\sigma = \frac{p_S}{\mu_I}, \tag{14}$$

$$\hat{\nu} = \nu p_T \ \text{(they always appear together)}, \tag{15}$$

$$\xi(\hat{\nu}) = \left(1 + \frac{\hat{\nu}}{\mu_I}\right)^{-1}, \tag{16}$$

$$\psi = \frac{h_T}{\mu_T}. \tag{17}$$

Using the values in Table S1, we get $\omega = 9.59\,hour$, $\sigma = 46.63\,hour$, $\psi = 22.51\,hour$. We assume $p_1 = p_2 = p$, and write the `ZIP` equations for the effective SIS parameters at the critical point (Eq. (7)):

$$\left\{\lambda^{(w)}_{critical}\,\rho[\bar{A}]\right\}^{-1} = \omega + \sigma\xi(\hat{\nu})\left[1 + \psi(1-p)^2\hat{\nu}\right]; \tag{18}$$

$$\left\{\lambda^{(1)}_{critical}\,\rho[\bar{A}]\right\}^{-1} = (1-\phi_1)\left\{\omega + \sigma\xi(\hat{\nu})\left[1 + \psi(1-p)\hat{\nu}\right]\right\}; \tag{19}$$

$$\left\{\lambda^{(12)}_{critical}\,\rho[\bar{A}]\right\}^{-1} = (1-\phi_1)(1-\phi_2)\left(\omega + \sigma\right). \tag{20}$$

From finding the minimum among Eq. (18,19,20), we recover the parameter surfaces that separate the different dominance phases (Fig. 4d of main paper). The network component $\rho[\bar{A}]$ always factors, so that network topology does not influence dominance [6] if the weak-commutation condition holds. For these reasons the following quantities are valid for any cntact network:

$$\hat{\nu}_{w-12} = \mu_I \frac{(\omega+\sigma)(\phi_1+\phi_2-\phi_1\phi_2)}{(1-\phi_1)(1-\phi_2)\sigma - \omega(\phi_1+\phi_2-\phi_1\phi_2) - p_S\frac{h_T}{\mu_T}(1-p)^2}; \quad (21)$$

$$\hat{\nu}_{w-1} = \frac{(\omega+\sigma)(1-\phi_1)}{\sigma\psi(1-p)(p-\phi_1) - \frac{\omega}{\mu_I}\phi_1}; \quad (22)$$

$$\hat{\nu}_{1-12} = \frac{-\phi_2(\omega+\sigma)}{\sigma\psi(1-p) + \frac{\omega-(1-\phi_2)(\omega+\sigma)}{\mu_I}}. \quad (23)$$

We also define

$$g = \sigma\frac{1-\mu_I(1-p)^2\psi}{\omega+\sigma}; \quad (24)$$

$$\hat{g} = \frac{\psi p(1-p)}{\psi(1-p)+\omega/p_S}; \quad (25)$$

$$\tilde{g} = \sigma\frac{1-\mu_I(1-p)\psi}{\omega+\sigma}. \quad (26)$$

We then have that if $\phi_1+\phi_2-\phi_1\phi_2 > g$, then always $\lambda^{(w)}_{critical} < \lambda^{(12)}_{critical}$. If $0 < \phi_1+\phi_2-\phi_1\phi_2 < g$, then $\lambda^{(w)}_{critical} < \lambda^{(12)}_{critical} \Leftrightarrow \hat{\nu} < \hat{\nu}_{w-12}$, and $\lambda^{(w)}_{critical} > \lambda^{(12)}_{critical} \Leftrightarrow \hat{\nu} > \hat{\nu}_{w-12}$. Finally, if $\phi_1+\phi_2-\phi_1\phi_2 < 0$, then always $\lambda^{(w)}_{critical} > \lambda^{(12)}_{critical}$.

We then have that if $\phi_1 > \hat{g}$, then always $\lambda^{(w)}_{critical} < \lambda^{(1)}_{critical}$. If $0 < \phi_1 < \hat{g}$, then $\lambda^{(w)}_{critical} < \lambda^{(1)}_{critical} \Leftrightarrow \hat{\nu} < \hat{\nu}_{w-1}$, and $\lambda^{(w)}_{critical} > \lambda^{(1)}_{critical} \Leftrightarrow \hat{\nu} > \hat{\nu}_{w-1}$. Finally, if $\phi_1 < 0$, then always $\lambda^{(w)}_{critical} > \lambda^{(1)}_{critical}$.

We then have that if $\phi_2 > \tilde{g}$, then always $\lambda^{(1)}_{critical} < \lambda^{(12)}_{critical}$. If $0 < \phi_2 < \tilde{g}$, then $\lambda^{(1)}_{critical} < \lambda^{(12)}_{critical} \Leftrightarrow \hat{\nu} < \hat{\nu}_{1-12}$, and $\lambda^{(1)}_{critical} > \lambda^{(12)}_{critical} \Leftrightarrow \hat{\nu} > \hat{\nu}_{1-12}$. Finally, if $\phi_2 < 0$, then always $\lambda^{(1)}_{critical} > \lambda^{(12)}_{critical}$.

We can rewrite the last if we fix $\phi = \phi_1+\phi_2-\phi_1\phi_2$, and express as a function of $\phi_1$. We then have that if $\phi_1 < (\phi-\tilde{g})/(1-\tilde{g})$, then always $\lambda^{(1)}_{critical} < \lambda^{(12)}_{critical}$. If $(\phi-\tilde{g})/(1-\tilde{g}) < \phi_1 < \phi$, then $\lambda^{(1)}_{critical} < \lambda^{(12)}_{critical} \Leftrightarrow \hat{\nu} < \hat{\nu}_{1-12}$, and $\lambda^{(1)}_{critical} > \lambda^{(12)}_{critical} \Leftrightarrow \hat{\nu} > \hat{\nu}_{1-12}$. Finally, if $\phi_1 > \phi$, then always $\lambda^{(1)}_{critical} > \lambda^{(12)}_{critical}$.

The values of $g, \hat{g}, \tilde{g}$ are in Table S4, which shows that typically $\hat{g}$ is small. This means that is difficult – but not impossible – to get to the dominance of the one-resistance form if neither one-resistant strain has an advantage in terms of transmission.

| | $g$ | $\hat{g}$ | $\tilde{g}$ |
|---|---|---|---|
| $p = 10^{-1}$ | 0.6122 | 0.0586 | 0.5880 |
| $p = 10^{-2}$ | 0.5665 | 0.0061 | 0.5639 |
| $p = 10^{-3}$ | 0.5617 | 0.0006 | 0.5615 |

Table S4: Numerical values of $g, \hat{g}, \tilde{g}$, defined in Eq. (24,25,26), and calculated with the parameters in Table S1.

#### 2.3 Additional parameters definition

Table S5 reports the definition of the additional parameters used in the rates of Fig. 4a of the main text. They are combinations of the parameters of the original model (Table S1).

| parameter | value | index value |
|---|---|---|
| $\alpha_x =$ | $(1-p_1)(1-p_2)$ | for $x = w$ |
| $=$ | $(1-p_2)$ | $x = 1$ |
| $=$ | $(1-p_1)$ | $x = 2$ |
| $\phi_x =$ | $0$ | for $x = w$ |
| $=$ | $\phi_1$ | $x = 1$ |
| $=$ | $\phi_2$ | $x = 2$ |
| $\bar{p}_x =$ | $1-(1-p_1)(1-p_2)$ | for $x = w$ |
| $=$ | $p_2$ | $x = 1$ |
| $=$ | $p_1$ | $x = 2$ |
| $\phi_y =$ | $\phi_x$ | for $y = x$ |
| $=$ | $1-(1-\phi_1)(1-\phi_2)$ | $y = 12$ |

Table S5: Definitions of the additional parameters used in Fig. 4a of the main text. The indices take the following values: $x = w, 1, 2$; $y = w, 1, 2, 12$, as in Fig 4a.

### 3 COVID-19

We model COVID-19 following Refs. [7, 8], using a notation similar to the one adopted to introduce the pandemic influenza case study. Once infected, susceptible individuals (S) enter the exposed compartment (E). After an average latency period $\epsilon^{-1}$ individuals become pre-symptomatic (P). A proportion $1 - p_S$ of them does not develop symptoms and enters compartment A at rate $\epsilon_P$. The other become paucisymptomatic ($I_{ps}$, prob $p_{ps}$), mildly symptomatic ($I_{ms}$, prob $p_{ms}$), or develop severe symptoms ($I_{ss}$, $1 - p_{ms} - p_{ss}$). $A, I_{ps}, I_{ms}, I_{ss}$ recover at rate $\mu$. $I_{ss}$

transition at the same rate to either hospitalization in intensive-care unit, ICU (U, prob $p_{ICU}$), or hospitalization outside ICU (H, prob $1 - p_{ICU}$). Both can either die or recover at specific rates. The compartmental scheme is shown in Figure S4.

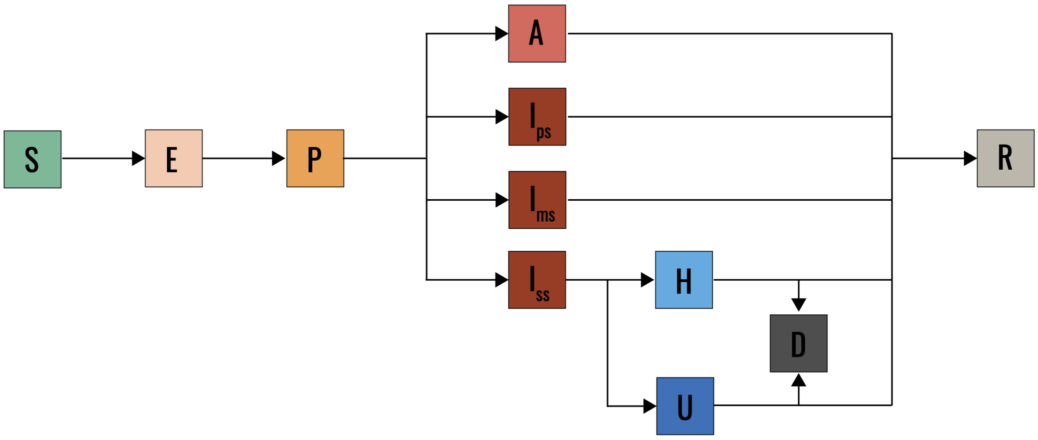


Figure S4: Compartmental model for COVID-19 [7, 8].

Compartments $\text{P}, \text{A}, \text{I}_{ps}, \text{I}_{ms}, \text{I}_{ss}$ can transmit SARS-CoV-2. Upon contact, $\text{P}, \text{A}, \text{I}_{ps}$ do so at rate $\lambda$, while $\text{I}_{ps}, \text{I}_{ms}$ at rate $\eta\lambda$, with $\eta = 0.51$ as in Refs. [7, 8].

A key aspect of this model is that transition probability depend on age. We consider four age classes (0-10; 11-18; 19-64; and 65+ years old, corresponding to children, adolescents, adults, and seniors), and the corresponding parameters are reported in Refs. [7,8]. Crucially, for instance, young individuals who develop symptoms are always paucisymptomatic ($p_{ps} = 1$), while $p_{ps} = 0.2$ for adults and seniors.

Practically this means parameters $p_{ps}, p_{ms}$ are $N \times N$ diagonal matrices where $i$th diagonal entry contains the value corresponding to the age class of the $i$th individual.

As for the influenza model, the COVID-19 model can be `CUT`, and `ZIP`ed if in timescale separation.

Figure S5 reports the model after `CUT`. The `ZIP`ing process is similar to influenza's, with the difference that some parameters are matrices. This leads to the following equation of the threshold:

$$\lambda_c = \frac{1}{\rho\left[\bar{A}\left\{\epsilon_p^{-1} + \mu^{-1}(1 - p_S) + \mu^{-1} p_S\left[\eta + (1 - \eta)p_{ps}\right]\right\}\right]}. \tag{27}$$

Below are the details of the `ZIP`ping.

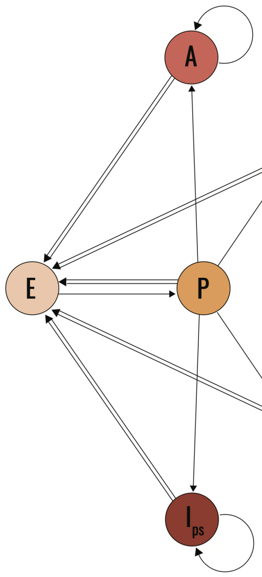


Figure S5: EGD of the COVID-19 model after `CUT`.

##### 3.1 `ZIP` of COVID-19

The Jacobian of the relevant subdiagram reads

$$J_X = \begin{pmatrix} -\epsilon_P & \epsilon_P(1-p_S) & \epsilon_P p_S p_{ps} & \epsilon_P p_S p_{ms} & \epsilon_P p_S p_{ss} \\ 0 & -\mu & 0 & 0 & 0 \\ 0 & 0 & -\mu & 0 & 0 \\ 0 & 0 & 0 & -\mu & 0 \\ 0 & 0 & 0 & 0 & -\mu \end{pmatrix}. \tag{28}$$

This can be inverted using block matrix inversion:

$$J_X^{-1} = \left( \begin{array}{c|c} -\epsilon_P^{-1} & -\mu^{-1} B \\ \hline 0 & \mu^{-1} \end{array} \right), \tag{29}$$

with

$$B = \begin{bmatrix} 1-p_S & p_S p_{ps} & p_S p_{ms} & p_S(1-p_{ps}-p_{ms}) \end{bmatrix}. \tag{30}$$

From this, and by using `ZIP` equations, one gets to Eq. 27.
